# Evaluating the association of *APOE* genotype and cognitive resilience in SuperAgers

**DOI:** 10.1101/2025.01.07.25320117

**Authors:** Alaina Durant, Shubhabrata Mukherjee, Michael L. Lee, Seo-Eun Choi, Phoebe Scollard, Brandon S. Klinedinst, Emily H. Trittschuh, Jesse Mez, Lindsay A. Farrer, Katherine A. Gifford, Carlos Cruchaga, Jason Hassenstab, Adam C. Naj, Li-San Wang, Sterling C. Johnson, Corinne D. Engelman, Walter A. Kukull, C. Dirk Keene, Andrew J. Saykin, Michael L. Cuccaro, Brian W. Kunkle, Margaret A. Pericak-Vance, Eden R. Martin, David A. Bennett, Lisa L. Barnes, Julie A. Schneider, William S. Bush, Jonathan L. Haines, Richard Mayeux, Badri N. Vardarajan, Marilyn S. Albert, Paul M. Thompson, Angela L. Jefferson, The Alzheimer’s Disease Neuroimaging Initiative (ADNI), Alzheimer’s Disease Genetics Consortium (ADGC), The Alzheimer’s Disease Sequencing Project (ADSP), Paul K. Crane, Logan Dumitrescu, Derek B. Archer, Timothy J. Hohman, Leslie S. Gaynor

## Abstract

**INTRODUCTION:** “SuperAgers” are oldest-old adults (ages 80+) whose memory performance more closely resembles middle-aged adults. The present study examined *APOE* allele frequency in non-Hispanic Black (NHB) and non-Hispanic White (NHW) SuperAgers compared to controls and Alzheimer’s disease dementia cases.

**METHODS:** In 18,080 participants from eight cohorts, harmonized clinical diagnostics and memory, executive function, and language domain scores were used to identify SuperAgers, cases, and controls across age-defined bins.

**RESULTS:** NHW SuperAgers had significantly lower frequency of *APOE-*ε4 alleles and higher frequency of *APOE*-ε2 alleles compared to all cases and controls, including oldest-old controls. Similar patterns were found in a small yet substantial sample of NHB SuperAgers; however, not all comparisons with controls reached significance.

**DISCUSSION:** We demonstrated strong evidence that *APOE* allele frequency relates to SuperAger status. Further research is needed with a larger sample of NHB SuperAgers to determine if mechanisms conferring resilience differ across race groups.

## 1 Background

“SuperAgers” is a term used to describe oldest-old (ages 80+) adults with episodic memory performance most closely resembling adults in their 50s to mid-60s.^1,2^ Studies suggest that resilience to Alzheimer’s disease (AD) pathological changes and neurodegeneration may explain SuperAgers’ high memory scores.^3–6^ Further research is needed to elucidate factors conferring resilience to AD-related brain changes and subsequent cognitive decline in SuperAgers. Moreover, research is needed to explore resiliency factors in non-Hispanic Black (NHB) SuperAgers, as this group is largely understudied.^7^

*APOE-*ε4 is the strongest genetic risk factor for late-onset AD.^8^ The Northwestern SuperAging project reported lower *APOE-*ε4 allele frequency in SuperAgers (N = 10-12) compared to non-demented older adults.^2,9^ In contrast, most studies report no differences in *APOE*-ε4 allele frequency between SuperAgers and oldest-old adults with typical memory performance, both groups having lower *APOE*-ε4 allele frequency compared to AD dementia cases.^4–6,10–12^ Notably, these studies have small SuperAger samples (N = 25-64)^4–6,10–12^ oftentimes drawn from the same cohort, thus limiting their generalizability and reliability. Further, these studies exclusively include NHW participants.^4–6,10–12^ To our knowledge, only one study has been published with a NHB SuperAger sample (N = 61) and did not find a significant difference in *APOE*-ε4 allele frequency between SuperAgers and same-age controls.^7^ Even fewer studies have explored the relationship of *APOE*-ε2, the protective *APOE* allele, and SuperAger status,^11–13^ likely due to the low minor allele frequency of *APOE*-ε2. Studies of *APOE*-ε2 allele frequency and superior memory in the oldest-old have not found a significant relationship^11–13^; however, questions of statistical power, generalizability, and reliability remain.

The present study aims to explore *APOE*-ε4 and -ε2 allele frequency in SuperAgers compared to AD dementia cases and controls in a large, harmonized multicohort dataset from the Alzheimer’s Disease Sequencing Project Phenotype Harmonization Consortium (ADSP-PHC). Using harmonized clinical diagnoses and cognitive domain scores (e.g., memory, executive function, language), we classified NHW and NHB middle-aged, old, and oldest-old adults as cases, controls, or SuperAgers, and compared *APOE*-ε4 and -ε2 allele frequency of SuperAgers to cases and controls by age bin. Although prior literature suggests that there is not a relationship between optimal memory in oldest-old age and *APOE* genotype, this is likely due to a limitation of sample size. The ADSP-PHC has enabled us to complete, to our knowledge, the largest and most racially diverse study to date of *APOE* allele frequency and SuperAger status. We hypothesize that SuperAgers will possess a lower frequency of *APOE*-ε4 alleles and a higher frequency of *APOE*-ε2 alleles compared to both AD dementia cases and controls.

## 2 Methods

### 2.1 Study Population

The ADSP-PHC was assembled in 2021 to provide large-scale harmonization of ADSP cohorts, spanning markers of cognition, neuroimaging, fluid biomarkers, and neuropathology. Cohorts that are part of ADSP-PHC and were included in the present study are: Adult Changes in Thought (ACT),^14^ Alzheimer’s Disease Neuroimaging Initiative (ADNI),^15^ Biomarkers of Cognitive Decline Among Normal Individuals (BIOCARD),^16^ National Alzheimer’s Coordinating Centers (NACC),^17^ National Institute on Aging Alzheimer’s Disease Family Based Study (NIA-AD FBS),^18^ Religious Orders Study/Rush Memory and Aging Project/Minority Aging Research Study (ROS/MAP/MARS),^19,20,21^ Knight Alzheimer’s Disease Research Center (Knight ADRC),^22^ and Wisconsin Registry for Alzheimer’s Prevention (WRAP).^23^

Written informed consent was obtained from all participants in each cohort, and research was carried out with protocols approved by each site’s institutional review board. These secondary analyses were approved by the Vanderbilt University Medical Center institutional review board.

### 2.2 Cognitive Domain Scores

Each cohort used different neuropsychological assessment tools to measure cognition which may not be on the same scale. We derived co-calibrated and harmonized cognitive domain scores for four domains (i.e., memory, executive function, language, and visuospatial) using confirmatory factor analysis.^24^ Consistent with previous definitions of SuperAgers, only memory, executive function, and language domains were used in present analyses.^1,2^ To ensure inclusion of high-quality harmonization, cognitive domain scores with a standard error of measurement > 0.6 (estimated during the co-calibration and composite generation procedure) were excluded. Visit the data dictionary at https://vmacdata.org/adsp-phc/data/data-dictionary/cognition to explore the cognitive variables from each cohort that make up each cognitive domain.

For the purpose of this study, regression-based cognitive domain scores were created for participant classification. Age, years of education, sex, and cohort were regressed on cognitive domain score at each available timepoint. Residuals were used to capture cognitive performance not explained by demographic variables or cohort differences. Separate models were used for NHW and NHB participants.

### 2.3 Participant Classification

Participants were grouped by race (self or study reported), clinical status, and age (**Figure 1A**). Age bins were decided based on the age-related criteria of SuperAgers (ages 80+) and the middle-aged adults whose memory performance is compared to oldest-old adults to determine SuperAger status (ages 50-64).^25^ Therefore, the age bins included middle-aged (ages 50-64), old (ages 65-79), and oldest-old (ages 80+) adults. Participant classification was determined separately for NHW and NHB adults. Given that longitudinal data were available for most participants, and some participants’ ages spanned multiple age bins, participant classification was decided using a predetermined schema (**Figure 1B**).

**Figure 1.**
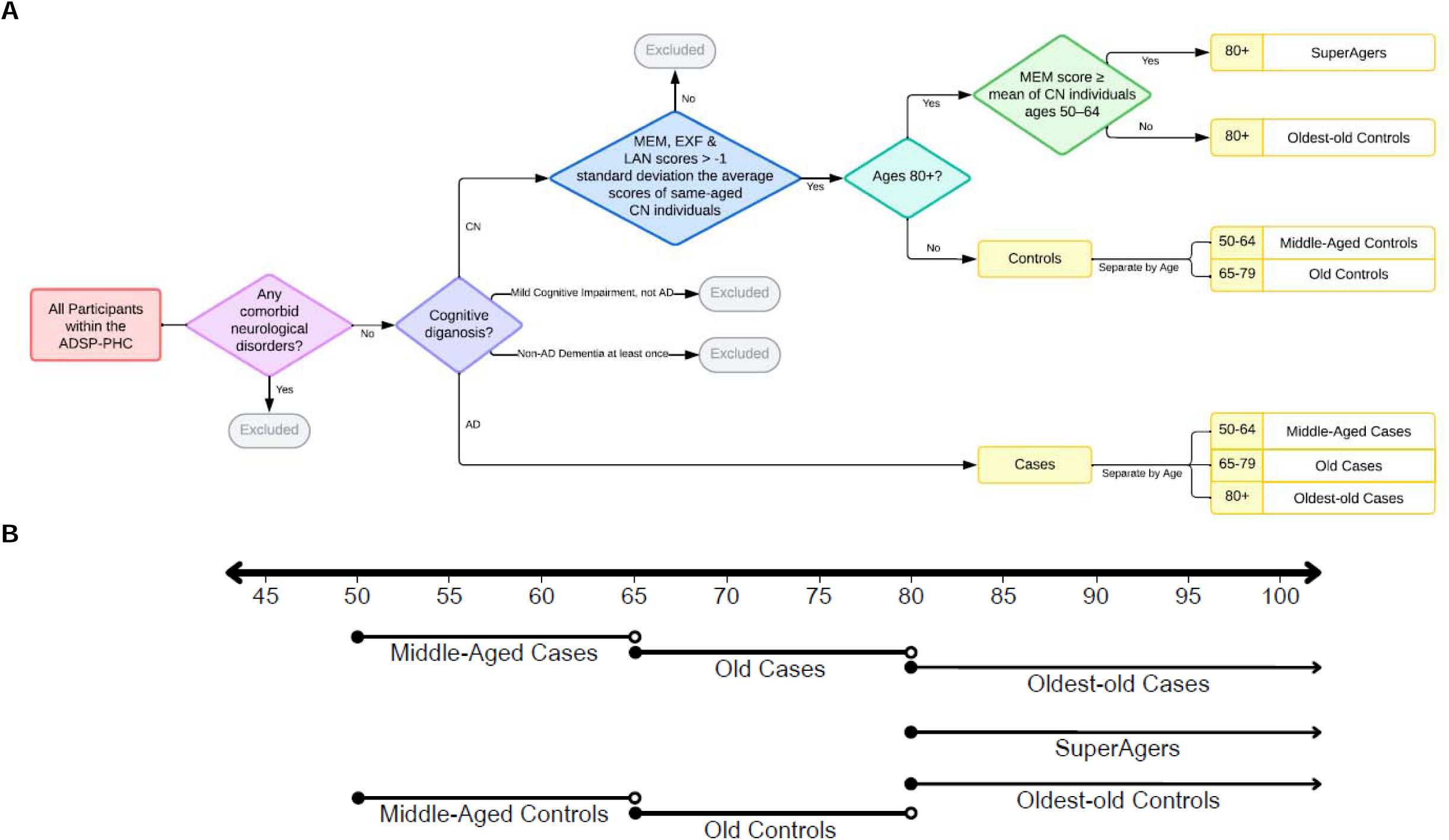
Flow Diagram for Participant Classification of SuperAgers, Cases, and Controls. (A) Flowchart depicting inclusion and exclusion criteria for identifying SuperAgers, AD dementia cases, controls. (B) Flowchart depicting selection order of SuperAgers, cases, and controls. Age range of participants indicated by line segment with arrows on each end. Age of participant classification is indicated by position of shorter, labeled line segments. Closed circles at the end of line segments indicate inclusion of age, such that age range is less-than-or-equal-to or greater-than-or-equal-to the age with which the circle aligns, while open circles indicate exclusion of age, such that age range is less-than or greater-than the age with which the circle aligns. Sequence of selection is indicated by line height, higher lines indicating earlier selection. Abbreviations: ADSP-PHC, Alzheimer’s Disease Sequencing Project – Phenotype Harmonization Consortium; AD, Alzheimer’s Disease; CN, Cognitively Normal; MEM, Memory; EXF, Executive Functioning; LAN, Language.

Firstly, participants with a clinical diagnosis of AD dementia at least once were considered cases. Across all cohorts, a diagnosis of AD dementia followed standard published criteria (National Institute of Neurological and Communicative Disorders and Stroke and the Alzheimer’s Disease and Related Disorders Association, or NINCDS-ADRDA).^26–28^ Given that the present study focused on dementia cases likely due to AD, participants with comorbid neurological disorders (e.g., stroke, traumatic brain injury with significant loss of consciousness) or primary diagnoses of non-AD dementia (e.g., Dementia with Lewy Bodies, Vascular Dementia) at any timepoint were excluded. Age bin was determined based on age at first AD dementia diagnosis.

Next, SuperAgers were identified using previously published criteria.^1,2^ Across all cohorts, SuperAgers were defined as oldest-old adults with (1) memory scores at or above the mean of middle-aged adults (ages 50-64), (2) executive function and language residual scores no more than 1 standard deviation below their same-aged peers (ages 80+) at the same visit as their superior memory performance, and (3) whose diagnosis remained “cognitively normal” for the duration of study participation.

Regarding classification as a control, given the importance of oldest-old (ages 80+) participants to our central analyses, we sequentially identified oldest-old, middle-aged, and then old controls. Criteria for controls included (1) memory, executive function, and language residual scores no lower than 1 standard deviation below their same-aged peers at a single visit, and (2) whose diagnosis remained “cognitively normal” for the duration of their study participation.

### 2.4 *APOE* Genotyping

*APOE* haplotypes were determined from the single nucleotide variants rs7412 and rs429358 for ACT, BIOCARD, NACC, NIA-AD FBS, Knight ADRC, and WRAP and from pyrosequencing of *APOE* codons 112 and 158 for ADNI and ROS/MAP/MARS.^29,30^

### 2.5 Statistical Analyses

Statistical analyses were performed using R Statistical Software (v4.2.3.).^31^ Logistic regression models examined differences in *APOE*-ε4 and -ε2 allele frequency of SuperAgers compared to cases and controls at all age bins*. APOE* allele positivity was determined by allele presence (0 = no allele present, 1 = one or more alleles present). Models covaried for sex and years of education due to their known modifying effects on the relationship of *APOE* and late-life cognition.^32,33^ Sensitivity analyses included analyses removing individuals with *APOE*-ε2/ε4 genotype and adding *APOE* genotyping method as a covariate. Additionally, to determine whether differences in sample size explained differences in the significance of comparisons across NHW and NHB samples, we included sensitivity analyses randomly down-sampling the NHW sample by participant classification and cohort. Correction for multiple comparisons was applied using Benjamini-Hochberg false discovery rate (FDR) procedure.^34^ Results tables include odds ratios (OR), confidence intervals (CI), and FDR-corrected *p*-values.

## 3 Results

### 3.1 Participant Characteristics

In total, 18,080 participants were included in the present analyses with a total of 78,549 datapoints (**Table 1**). Participants completed an average of 4 ± 4 visits over 5 ± 5 years. The number of follow-up visits and length of follow-up varied by cohort due to differences in study design.

**Table 1.**
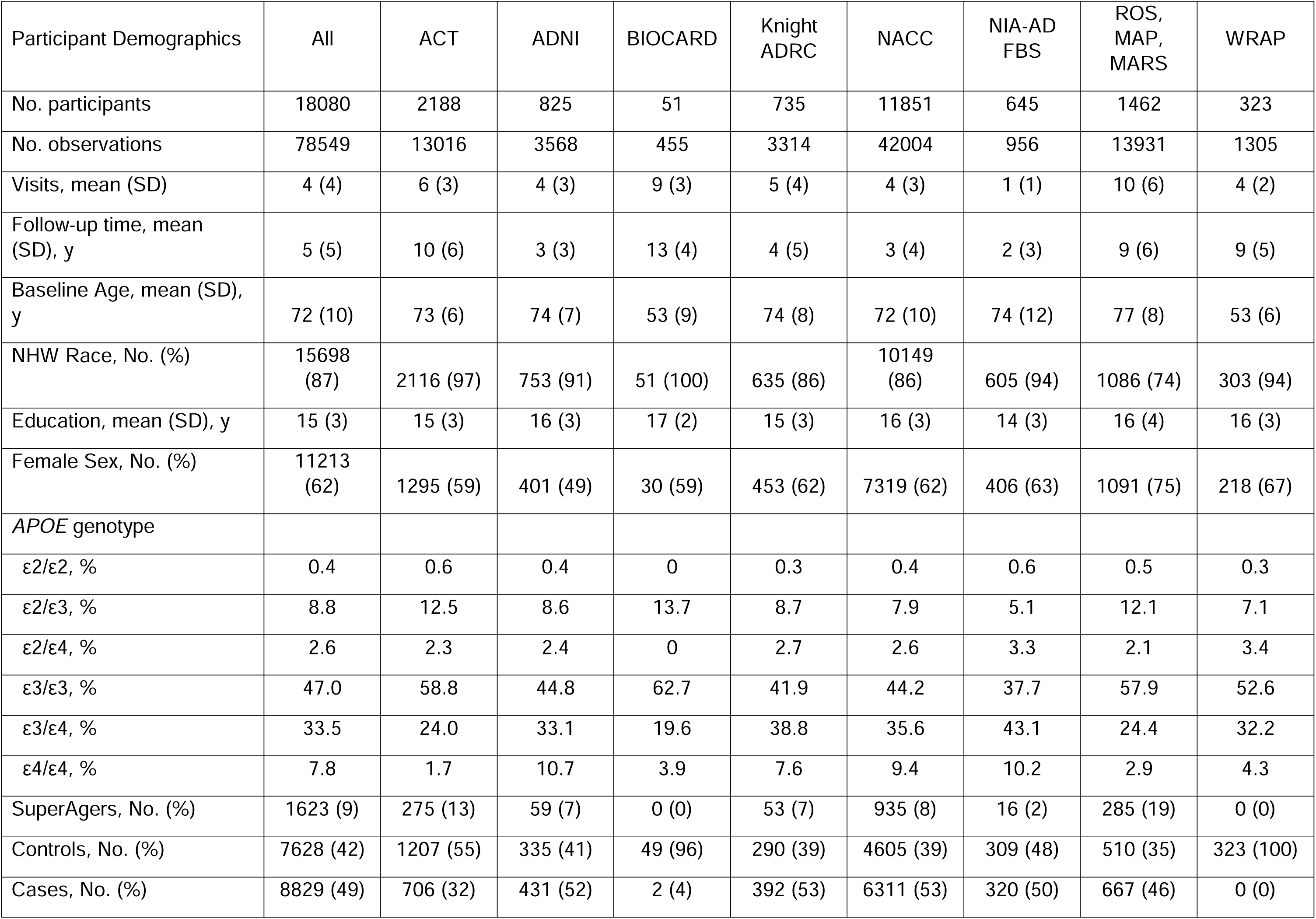
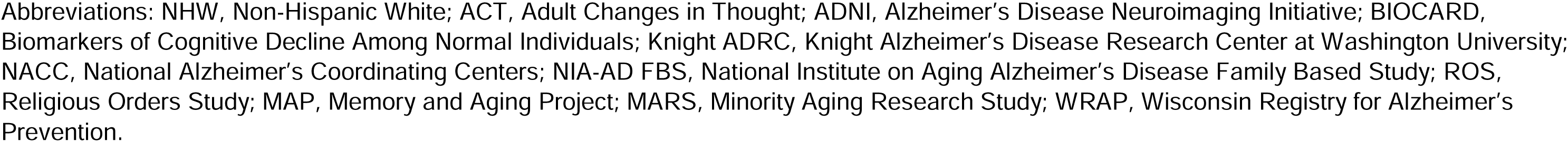
Participant Characteristics by Cohort.

| Participant Demographics | All | ACT | ADNI | BIOCARD | Knight<br>ADRC | NACC | NIA-AD<br>FBS | ROS,<br>MAP,<br>MARS | WRAP |
| --- | --- | --- | --- | --- | --- | --- | --- | --- | --- |
| No. participants | 18080 | 2188 | 825 | 51 | 735 | 11851 | 645 | 1462 | 323 |
| No. observations | 78549 | 13016 | 3568 | 455 | 3314 | 42004 | 956 | 13931 | 1305 |
| Visits, mean (SD) | 4 (4) | 6 (3) | 4 (3) | 9 (3) | 5 (4) | 4 (3) | 1 (1) | 10 (6) | 4 (2) |
| Follow-up time, mean<br>(SD), y | 5 (5) | 10 (6) | 3 (3) | 13 (4) | 4 (5) | 3 (4) | 2 (3) | 9 (6) | 9 (5) |
| Baseline Age, mean (SD),<br>y | 72 (10) | 73 (6) | 74 (7) | 53 (9) | 74 (8) | 72 (10) | 74 (12) | 77 (8) | 53 (6) |
| NHW Race, No. (%) | 15698<br>(87) | 2116 (97) | 753 (91) | 51 (100) | 635 (86) | 10149<br>(86) | 605 (94) | 1086 (74) | 303 (94) |
| Education, mean (SD), y | 15 (3) | 15 (3) | 16 (3) | 17 (2) | 15 (3) | 16 (3) | 14 (3) | 16 (4) | 16 (3) |
| Female Sex, No. (%) | 11213<br>(62) | 1295 (59) | 401 (49) | 30 (59) | 453 (62) | 7319 (62) | 406 (63) | 1091 (75) | 218 (67) |
| *APOE* genotype |  |  |  |  |  |  |  |  |  |
| $\epsilon 2/\epsilon 2$, % | 0.4 | 0.6 | 0.4 | 0 | 0.3 | 0.4 | 0.6 | 0.5 | 0.3 |
| $\epsilon 2/\epsilon 3$, % | 8.8 | 12.5 | 8.6 | 13.7 | 8.7 | 7.9 | 5.1 | 12.1 | 7.1 |
| $\epsilon 2/\epsilon 4$, % | 2.6 | 2.3 | 2.4 | 0 | 2.7 | 2.6 | 3.3 | 2.1 | 3.4 |
| $\epsilon 3/\epsilon 3$, % | 47.0 | 58.8 | 44.8 | 62.7 | 41.9 | 44.2 | 37.7 | 57.9 | 52.6 |
| $\epsilon 3/\epsilon 4$, % | 33.5 | 24.0 | 33.1 | 19.6 | 38.8 | 35.6 | 43.1 | 24.4 | 32.2 |
| $\epsilon 4/\epsilon 4$, % | 7.8 | 1.7 | 10.7 | 3.9 | 7.6 | 9.4 | 10.2 | 2.9 | 4.3 |
| SuperAgers, No. (%) | 1623 (9) | 275 (13) | 59 (7) | 0 (0) | 53 (7) | 935 (8) | 16 (2) | 285 (19) | 0 (0) |
| Controls, No. (%) | 7628 (42) | 1207 (55) | 335 (41) | 49 (96) | 290 (39) | 4605 (39) | 309 (48) | 510 (35) | 323 (100) |
| Cases, No. (%) | 8829 (49) | 706 (32) | 431 (52) | 2 (4) | 392 (53) | 6311 (53) | 320 (50) | 667 (46) | 0 (0) |

Abbreviations: NHW, Non-Hispanic White; ACT, Adult Changes in Thought; ADNI, Alzheimer's Disease Neuroimaging Initiative; BIOCARD, Biomarkers of Cognitive Decline Among Normal Individuals; Knight ADRC, Knight Alzheimer's Disease Research Center at Washington University; NACC, National Alzheimer's Coordinating Centers; NIA-AD FBS, National Institute on Aging Alzheimer's Disease Family Based Study; ROS, Religious Orders Study; MAP, Memory and Aging Project; MARS, Minority Aging Research Study; WRAP, Wisconsin Registry for Alzheimer's Prevention.

Average baseline age varied by cohort (Age_[all cohorts]_ = 72 ± 10); the youngest cohorts on average, BIOCARD and WRAP, primarily recruited cognitively normal participants. Generally, cohorts were highly educated (Years of Education_[all cohorts]_ = 15 ± 3), mostly female (62.9%), and mostly NHW (85.4%). The frequency of each *APOE* genotype differed by cohort, with higher frequency of ε3/ε4 and ε4/ε4 genotypes in cohorts with a higher frequency of AD dementia cases.

### 3.2 Participant Classification

SuperAgers made up 9% of all participants (N = 1,623). The two youngest cohorts, BIOCARD and WRAP, did not contribute any SuperAgers. Cognitively unimpaired controls comprised 42% of all participants (N = 7,628), and cases made up 49% of all participants (N = 8,829).

**Table 2** displays participant characteristics of NHW and NHB SuperAgers, controls, and cases in age-defined bins. On average, NHW SuperAgers (N = 1,412) were somewhat older, had more years of education, and included more males than NHB SuperAgers (N = 211). There was not a difference in the proportion of participants in each participant classification (SuperAger, control, case) across racialized groups (χ^2^_[2]_ = 3.79, p = 0.15). Comparing NHW and NHB participants across age-defined bins, all NHW bins had greater average years of education and a higher proportion of males than NHB bins.

**Table 2.** Characteristics of SuperAgers, Cases, and Controls by Race and Age Bin. A.

|  | SuperAgers | Middle-Aged Controls | Old Controls | Oldest-old Controls | Middle-Aged Cases | Old Cases | Oldest-old Cases |
| --- | --- | --- | --- | --- | --- | --- | --- |
| No. participants | 1412 | 1622 | 3202 | 1213 | 1101 | 3528 | 3258 |
| No. observations | 11528 | 7067 | 12308 | 6980 | 2692 | 10858 | 15434 |
| Visits, mean (SD) | 8 (5) | 4 (3) | 4 (3) | 6 (4) | 2 (2) | 3 (2) | 5 (4) |
| Follow-up time, mean (SD), y | 10 (6) | 6 (5) | 4 (4) | 8 (6) | 2 (2) | 2 (3) | 5 (6) |
| Baseline age, mean (SD), y | 77 (7) | 57 (5) | 70 (4) | 79 (7) | 59 (5) | 73 (4) | 83 (6) |
| Education, mean (SD), y | 16 (3) | 16 (2) | 16 (3) | 15 (3) | 15 (3) | 15 (3) | 15 (3) |
| Female Sex, No. (%) | 953 (67) | 1119 (69) | 1964 (61) | 661 (54) | 618 (56) | 1905 (54) | 1989 (61) |
| *APOE*-ε2 Frequency, No. (%) | 246 (17) | 216 (13) | 467 (15) | 179 (15) | 50 (5) | 184 (5) | 321 (10) |
| *APOE*-ε4 Frequency, No. (%) | 276 (20) | 647 (40) | 981 (31) | 270 (22) | 639 (58) | 2495 (71) | 1425 (44) |

**B**
|  | SuperAgers | Middle-Aged Controls | Old Controls | Oldest-old Controls | Middle-Aged Cases | Old Cases | Oldest-old Cases |
| --- | --- | --- | --- | --- | --- | --- | --- |
| No. participants | 211 | 296 | 752 | 145 | 85 | 413 | 444 |
| No. observations | 1830 | 1024 | 3106 | 827 | 171 | 1300 | 2281 |
| Visits, mean (SD) | 9 (5) | 3 (3) | 4 (3) | 6 (4) | 2 (1) | 3 (3) | 5 (5) |
| Follow-up time, mean (SD), y | 9 (6) | 4 (4) | 4 (4) | 6 (5) | 1 (1) | 3 (3) | 5 (6) |
| Baseline age, mean (SD), y | 77 (6) | 59 (4) | 70 (4) | 79 (6) | 60 (4) | 73 (4) | 83 (6) |
| Education, mean (SD), y | 15 (3) | 15 (3) | 15 (3) | 13 (3) | 14 (3) | 14 (4) | 13 (4) |
| Female Sex, No. (%) | 177 (84) | 222 (75) | 564 (75) | 115 (79) | 58 (68) | 283 (69) | 331 (75) |
| *APOE*-ε2 Frequency, No. (%) | 56 (27) | 68 (23) | 155 (21) | 27 (19) | 8 (9) | 36 (9) | 53 (12) |
| *APOE*-ε4 Frequency, No. (%) | 54 (26) | 125 (42) | 261 (35) | 39 (27) | 65 (76) | 307 (74) | 216 (49) |

(A) Non-Hispanic White; (B) Non-Hispanic Black

### 3.3 *APOE* Allele Frequency

In NHW comparisons (**Table 3A**), SuperAgers had a significantly higher frequency of *APOE*-ε2 alleles (**Figure 2A**; middle-aged controls: OR = 1.38, 95%CI[1.13, 1.68], *P_FDR_* = 0.002; old controls: OR = 1.24, 95%CI[1.05, 1.47], *P_FDR_* = 0.015; oldest-old controls: OR = 1.28, 95%CI[1.03, 1.59], *P_FDR_* = 0.035; middle-aged cases: OR = 4.55, 95%CI[3.30, 6.27], *P_FDR_* < 0.001; old cases: OR = 4.02, 95%CI[3.26, 4.96], *P_FDR_* < 0.001; oldest-old cases: OR = 2.03, 95%CI[1.68, 2.44], *P_FDR_* < 0.001) and a significantly lower frequency of *APOE*-ε4 alleles (**Figure 2B**; middle-aged controls: OR = 0.37, 95%CI[0.31, 0.43], *P_FDR_* < 0.001; old controls: OR = 0.55, 95%CI[0.47, 0.64], *P_FDR_* < 0.001; oldest-old controls: OR = 0.81, 95%CI[0.67, 0.99], *P_FDR_* = 0.044; middle-aged cases: OR = 0.18, 95%CI[0.15, 0.21], *P_FDR_* < 0.001; old cases: OR = 0.09, 95%CI[0.08, 0.11], *P_FDR_* < 0.001; oldest-old cases: OR = 0.32, 95%CI[0.27,0.37], *P_FDR_* < 0.001) compared to all cases and controls. In contrast, NHB SuperAgers (**Table 3B**) had a significantly higher frequency of *APOE*-ε2 alleles only compared to cases (**Figure 2C**; middle-aged cases: OR = 3.59, 95%CI[1.60, 8.06], *P_FDR_* = 0.005; old cases: OR = 4.56, 95%CI[2.75, 7.55], *P_FDR_* < 0.001; oldest-old cases: OR = 2.73, 95%CI[1.75, 4.25], *P_FDR_* < 0.001), and a significantly lower frequency of *APOE*-ε4 alleles compared to all cases and controls except oldest-old controls (**Figure 2D**; middle-aged controls: OR = 0.48, 95%CI[0.33, 0.71], *P_FDR_* = 0.001; old controls: OR = 0.66, 95%CI[0.47, 0.94], *P_FDR_* = 0.035; middle-aged cases: OR = 0.12, 95%CI[0.07, 0.22], *P_FDR_* < 0.001; old cases: OR = 0.12, 95%CI[0.08, 0.18], *P_FDR_* < 0.001; oldest-old cases: OR = 0.39, 95%CI[0.27, 0.57], *P_FDR_* < 0.001).

**Table 3.** Logistic Regression Model Results Comparing *APOE-*ε2 and -ε4 Allele Frequency among SuperAgers, Cases, and Controls. A.

|  | APOE-ε2 |  | APOE-ε4 |  |
| --- | --- | --- | --- | --- |
| | OR (CI) | $P_{FDR}$ | OR (CI) | $P_{FDR}$ |
| SuperAgers vs. Middle-Aged Controls | 1.38 (1.13, 1.68) | 0.002 | 0.37 (0.31, 0.43) | <0.001 |
| SuperAgers vs. Old Controls | 1.24 (1.05, 1.47) | 0.015 | 0.55 (0.47, 0.64) | <0.001 |
| SuperAgers vs. Oldest-Old Controls | 1.28 (1.03, 1.59) | 0.035 | 0.81 (0.67, 0.99) | 0.044 |
| SuperAgers vs. Middle-Aged Cases | 4.55 (3.30, 6.27) | <0.001 | 0.18 (0.15, 0.21) | <0.001 |
| SuperAgers vs. Old Cases | 4.02 (3.26, 4.96) | <0.001 | 0.09 (0.08, 0.11) | <0.001 |
| SuperAgers vs. Oldest-Old Cases | 2.03 (1.68, 2.44) | <0.001 | 0.32 (0.27, 0.37) | <0.001 |

**B**
|  | APOE-ε2 |  | APOE-ε4 |  |
| --- | --- | --- | --- | --- |
| | OR (CI) | $P_{FDR}$ | OR (CI) | $P_{FDR}$ |
| SuperAgers vs. Middle-Aged Controls | 1.19 (0.79, 1.80) | 0.423 | 0.48 (0.33, 0.71) | 0.001 |
| SuperAgers vs. Old Controls | 1.41 (0.99, 2.01) | 0.079 | 0.66 (0.47, 0.94) | 0.035 |
| SuperAgers vs. Oldest-Old Controls | 1.63 (0.93, 2.85) | 0.115 | 1.18 (0.70, 1.98) | 0.564 |
| SuperAgers vs. Middle-Aged Cases | 3.59 (1.60, 8.06) | 0.005 | 0.12 (0.07, 0.22) | <0.001 |
| SuperAgers vs. Old Cases | 4.56 (2.75,<br>7.55) | $<0.001$ | 0.12 (0.08,<br>0.18) | $<0.001$ |
| SuperAgers vs. Oldest-Old Cases | 2.73 (1.75,<br>4.25) | $<0.001$ | 0.39 (0.27,<br>0.57) | $<0.001$ |

(A) Non-Hispanic White; (B) Non-Hispanic Black. Abbreviations: OR, Odds Ratio; CI, Confidence Interval (95%); $P_{FDR}$ , FDR-corrected P-value.

### 3.4 Sensitivity Analyses

Analyses were repeated removing individuals with an *APOE*-ε2/ε4 genotype. Given their low frequency, very few participants were removed (**Supplementary Table 1;** NHW SuperAgers: N = 1,393; NHB SuperAgers: N = 200) and findings were preserved (**Supplementary Table 2**).

Findings were also preserved when *APOE* genotyping method was included as a covariate (**Supplementary Table 3**).

To determine the impact of sample size on the significance of NHB comparisons, we randomly down-sampled the NHW sample by participant classification and cohort (**Supplementary Table 4**). In the smaller sample, differences in *APOE*-ε2 and -ε4 allele frequency between NHW SuperAgers and controls were no longer significant, except for the comparison between NHW SuperAgers and middle-aged controls (**Supplementary Table 5**).

## 4 Discussion

Mechanisms conferring resilience to memory decline in oldest-old age are yet unknown. This is the largest published sample of SuperAgers and the largest study to date of the relationship of *APOE*-ε4 and -ε2 allele frequency and SuperAger status. Across 8 national aging cohorts, we identified 1,623 NHW and NHB SuperAgers with *APOE* genotyping using longitudinal harmonized cognitive and clinical data. As expected, we found that SuperAgers had a higher frequency of *APOE*-ε2 alleles and a lower frequency of *APOE*-ε4 alleles compared to individuals with AD dementia. Unlike most previous studies of *APOE* genotype in NHW SuperAgers, we found significant differences in *APOE* genotype compared to controls of all ages, including oldest-old controls. Specifically, the odds of having 1 ε4 allele was about 20% lower in NHW SuperAgers compared to same-age controls, and the odds of having 1 ε2 allele was about 30% higher in NHW SuperAgers compared to same-age controls. This finding held when ε2/ε4 carriers were removed from analyses and when *APOE* genotyping method was added as a covariate. Although NHB SuperAgers also had a higher frequency of *APOE*-ε2 alleles and a lower frequency of *APOE*-ε4 alleles compared to oldest-old controls, comparisons did not rise to the level of significance. Sensitivity analyses down-sampling NHW SuperAgers suggest that sample size likely impacted the significance of comparisons with NHB SuperAgers.

*APOE*-ε4 is the strongest genetic risk factor for late-onset AD,^8^ and has been shown to be related to increased entorhinal and hippocampal atrophy^35^ and amnestic cognitive impairment.^36,37^ Our data are supportive of these findings, such that AD dementia cases had a higher frequency of *APOE*-ε4 alleles compared to controls and SuperAgers.^38^ Unlike most studies of genetic resilience in SuperAgers, we found that NHW SuperAgers had a lower frequency of *APOE*-ε4 alleles compared to oldest-old adults with typical memory performance. This was found in two previous studies published from the same cohort (N = 10-12),^2,9^ but was not replicated in subsequent studies with larger samples of SuperAgers.^4–6,10–12^ This finding is relatively unexpected. Across racialized groups, the effect of *APOE*-ε4 is most impressive prior to age 70.^39,40^ Additionally, *APOE*-ε4 carriership is related to increased mortality.^41,42^ In line with these studies, we found lower *APOE*-ε4 allele frequency in oldest-old compared to middle-aged and old controls. Despite NHW oldest-old controls being older than NHW SuperAgers on average, we found that SuperAgers had a significantly lower frequency of *APOE*-ε4 alleles compared to oldest-old controls, indicating that *APOE*-ε4 allele carriership influences memory even in adults who live past age 80. Moreover, dementia incidence increases exponentially with age,^43^ suggesting that the oldest-old control group also represents exceptional aging. We found that NHW SuperAgers had a significantly lower frequency of *APOE*-ε4 alleles compared to same-age peers, thus underscoring the utility of the SuperAgers phenotype for uncovering factors conferring resilience in cognitive aging.

The protective *APOE*-ε2 allele is related to lower likelihood of late-onset AD dementia^13,44^ and better cognitive performance in older adults even in the presence of AD neuropathology.^45^ Unlike *APOE*-ε4, *APOE*-ε2 carriership was previously shown to affect cognition after age 80; more precisely, although oldest-old *APOE*-ε2 carriers were as likely as *APOE*-ε4 carriers to meet neuropathologic criteria for AD, they were less likely to be diagnosed with dementia.^44^ Nonetheless, no previous studies have found a relationship of *APOE*-ε2 allele frequency and SuperAger status.^11–13^ The present study is the first to find that NHW SuperAgers had a significantly higher frequency of *APOE*-ε2 alleles compared to controls. Our results suggest that *APOE*-ε2 not only reduces the likelihood of dementia in oldest age but increases the likelihood one will possess optimal memory in oldest age. Future studies are needed to determine whether SuperAgers have similar levels of AD neuropathology compared to AD dementia cases as was previously found in oldest-old *APOE*-ε2 carriers without dementia.^44^

Genetic factors underlying the superior memory performance of NHB SuperAgers, a critically under-diagnosed and understudied group, are relatively unknown.^7^ Previous research suggests that, while NHB individuals have a higher frequency of *APOE*-ε4 alleles compared to NHW individuals, *APOE*-ε4 carriership is associated with attenuated risk for late-onset AD, yet similar mortality risk, in NHB individuals compared to NHW individuals.^40,46–48^ The effect is likely related to global population ancestry; more specifically, *APOE*-ε4 carriership is associated with a greater risk of AD in NHB older adults with decreased global African ancestry or increased global European ancestry.^40^ Similar to *APOE*-ε4, NHB individuals have a higher frequency of *APOE*-ε2 alleles compared to NHW individuals.^46^ Unlike *APOE*-ε4, researchers did not detect differences in the protective effect of *APOE*-ε2 alleles related to global population ancestry.^40^ In fact, a recent study from the MARS cohort found that NHB older adults (ages 65+) with more *APOE*-ε2 alleles had slower cognitive decline over a 10-year study period.^49^ Additionally, *APOE*-ε2 carriership was related to better survival in a sample of NHB and NHW individuals from NACC with and without AD neuropathology.^50^ In the present study, NHB SuperAgers had a significantly lower frequency of *APOE-*ε4 alleles compared to cases and younger controls, and a significantly higher frequency of *APOE-*ε2 alleles compared only to cases. Importantly, the present study included a substantially smaller sample of NHB SuperAgers compared to NHW SuperAgers, although the sample was still far larger than has been previously reported. Sensitivity analyses suggest that sample size likely impacted the significance of comparisons with NHB SuperAgers. It is also possible that the lack of significant difference in *APOE* allele carriership between NHB SuperAgers and oldest-old controls is partially explained by survivorship bias; recent research indicates that NHB adults have a higher probability of survival from ages 70 and 80 to 100 compared to NHW adults.^51^ Thus, NHB SuperAgers and cognitively normal oldest-old adults may share environmental and genetic factors, including *APOE-*ε2 allele carriership, that support survival and reduced mortality risk in older age, resulting in a lack of significant difference in *APOE* genotype between these two, very similar, exceptional aging groups.^7^ Still, more research and targeted recruitment of high-performing NHB oldest-old adults is necessary to determine the role of *APOE* genotype in their sustained optimal memory performance.

The present study has several strengths, including being the largest of its kind to explore the relationship of *APOE* genotype and optimal memory performance in both NHW and NHB SuperAgers. This is the largest study to date to identify differences in *APOE-*ε4 allele frequency based on SuperAger status, and the first study of SuperAgers to find a relationship between *APOE-*ε2 allele frequency and SuperAger status. Our study also has several limitations. Across cohorts, participants were highly educated, limiting generalizability to lower-educated or socioeconomically diverse populations. Additionally, studies have identified other genetic factors that may confer greater AD risk in NHB older adults compared to *APOE*, including *ABCA7*.^53,54^ Future studies will need to consider other genetic factors that may also be relevant to exceptional aging in NHB older adults. Moreover, differences in the effect of genetic factors on AD risk have been associated with genetic ancestry, which was only available for a subset of participants in our sample.^40^ Subsequent research leveraging advanced genetic analyses to consider admixture may further our understanding of genetic profiles underlying AD risk.

The present study reveals important information about genetic factors associated with SuperAger status. While significant findings were restricted to NHW comparisons, study results importantly direct our attention to the need for targeted recruitment of high-performing NHB oldest-old adults to more definitively determine the role of genetics in sustained optimal memory performance in oldest-old age across racialized groups. There is an ongoing initiative that intends to answer complex questions about brain aging, resilience, and resistance in a well-characterized SuperAging cohort through longitudinal, multimodal data collection.^55,56^ SuperAging research will contribute tremendously to our understanding of factors conferring resilience in oldest-old age, such as preserved regional brain volume, differences in lifestyle factors, and resistance to neuropathology.

## Data Availability

Data availability, deidentified participant data, and data dictionary can be accessed at the following url: https://www.vmacdata.org/adsp-phc. Corresponding data are available for request via the NIAGADS DSS.

https://vmacdata.org/adsp-phc/data/data-dictionary/cognition

## Acknowledgements/Conflicts/Funding Sources

The authors have nothing to disclose. The ADSP Phenotype Harmonization Consortium (ADSP-PHC) is funded by NIA (U24 AG074855, U01 AG068057 and R01 AG059716). This study was also funded by several additional funding sources, including K01 AG073584 (DBA) and K24 AG046373 (ALJ). The data contributed from the Adult Changes in Thought study was supported by NIA U19 AG066567. The data contributed from the Wisconsin Registry for Alzheimer’s Prevention was supported by NIA R01 AG021155, R01 AG0271761, R01 AG037639, and R01 AG054047. Data collection and sharing for this project was funded (in part) by the Alzheimer’s Disease Neuroimaging Initiative (ADNI) (NIA U01 AG024904) and DOD ADNI (Department of Defense award number W81XWH-12-2-0012). ADNI is funded by the National Institute on Aging, the National Institute of Biomedical Imaging and Bioengineering, and through generous contributions from the following: AbbVie, Alzheimer’s Association; Alzheimer’s Drug Discovery Foundation; Araclon Biotech; BioClinica, Inc.; Biogen; Bristol-Myers Squibb Company; CereSpir, Inc.; Cogstate; Eisai Inc.; Elan Pharmaceuticals, Inc.; Eli Lilly and Company; EuroImmun; F. Hoffmann-La Roche Ltd and its affiliated company Genentech, Inc.; Fujirebio; GE Healthcare; IXICO Ltd.; Janssen Alzheimer Immunotherapy Research & Development, LLC.; Johnson & Johnson Pharmaceutical Research & Development LLC.; Lumosity; Lundbeck; Merck & Co., Inc.; Meso Scale Diagnostics, LLC.; NeuroRx Research; Neurotrack Technologies; Novartis Pharmaceuticals Corporation; Pfizer Inc.; Piramal Imaging; Servier; Takeda Pharmaceutical Company; and Transition Therapeutics. The Canadian Institutes of Health Research is providing funds to support ADNI clinical sites in Canada. Private sector contributions are facilitated by the Foundation for the National Institutes of Health (www.fnih.org). The grantee organization is the Northern California Institute for Research and Education, and the study is coordinated by the Alzheimer’s Therapeutic Research Institute at the University of Southern California. ADNI data are disseminated by the Laboratory for Neuro Imaging at the University of Southern California. Data contributed from MAP/ROS/MARS was supported by NIA R01 AG017917, P30 AG10161, P30 AG072975, R01 AG022018, R01 AG056405, UH2 NS100599, UH3 NS100599, R01 AG064233, R01 AG15819 and R01 AG067482, and the Illinois Department of Public Health (Alzheimer’s Disease Research Fund). Data can be accessed at www.radc.rush.edu. The NACC database is funded by NIA/NIH Grant U24 AG072122. NACC data are contributed by the NIA-funded ADRCs: P30 AG062429 (PI James Brewer, MD, PhD), P30 AG066468 (PI Oscar Lopez, MD), P30 AG062421 (PI Bradley Hyman, MD, PhD), P30 AG066509 (PI Thomas Grabowski, MD), P30 AG066514 (PI Mary Sano, PhD), P30 AG066530 (PI Helena Chui, MD), P30 AG066507 (PI Marilyn Albert, PhD), P30 AG066444 (PI David Holtzman, MD), P30 AG066518 (PI Lisa Silbert, MD, MCR), P30 AG066512 (PI Thomas Wisniewski, MD), P30 AG066462 (PI Scott Small, MD), P30 AG072979 (PI David Wolk, MD), P30 AG072972 (PI Charles DeCarli, MD), P30 AG072976 (PI Andrew Saykin, PsyD), P30 AG072975 (PI Julie A. Schneider, MD, MS), P30 AG072978 (PI Ann McKee, MD), P30 AG072977 (PI Robert Vassar, PhD), P30 AG066519 (PI Frank LaFerla, PhD), P30 AG062677 (PI Ronald Petersen, MD, PhD), P30 AG079280 (PI Jessica Langbaum, PhD), P30 AG062422 (PI Gil Rabinovici, MD), P30 AG066511 (PI Allan Levey, MD, PhD), P30 AG072946 (PI Linda Van Eldik, PhD), P30 AG062715 (PI Sanjay Asthana, MD, FRCP), P30 AG072973 (PI Russell Swerdlow, MD), P30 AG066506 (PI Glenn Smith, PhD, ABPP), P30 AG066508 (PI Stephen Strittmatter, MD, PhD), P30 AG066515 (PI Victor Henderson, MD, MS), P30 AG072947 (PI Suzanne Craft, PhD), P30 AG072931 (PI Henry Paulson, MD, PhD), P30 AG066546 (PI Sudha Seshadri, MD), P30 AG086401 (PI Erik Roberson, MD, PhD), P30 AG086404 (PI Gary Rosenberg, MD), P20 AG068082 (PI Angela Jefferson, PhD), P30 AG072958 (PI Heather Whitson, MD), P30 AG072959 (PI James Leverenz, MD). NACC data can be accessed at naccdata.org. The recruitment and clinical characterization of research participants at Washington University were supported by NIH P30 AG066444 (JCM), P01 AG03991 (JCM), and P01 AG026276 (JCM). The National Institute on Aging-AD Family Based Study (NIA-AD FBS; https://www.neurology.columbia.edu/research/research-centers-and-programs/national-institute-aging-alzheimers-disease-family-based-study-nia-ad-fbs) collected the samples used in this study and is supported by National Institute on Aging (NIA) grant U24 AG056270. Additional families were contributed to the NIA-AD FBS through NIH grants: R01 AG028786, R01 AG027944, R01 AG027944, RF1 AG054074, U01 AG052410. This work was supported by access to equipment made possible by the Hope Center for Neurological Disorders, the Neurogenomics and Informatics Center (NGI: https://neurogenomics.wustl.edu/) and the Departments of Neurology and Psychiatry at Washington University School of Medicine. Data contributed from the Wisconsin Registry for Alzheimer’s Prevention were supported by R01 AG027161.

## Consent Statement

All participants provided informed consent in their respective cohort studies.

## Notes

### Competing Interest Statement

The authors have declared no competing interest.

### Author Declarations

IRB of Vanderbilt University Medical Center gave ethical approval for this work.

### Summary of Updates

Abstract and main text of the manuscript has minor revisions.

