## Supplementary Tables for "Evaluating the association of *APOE* genotype and cognitive resilience in SuperAgers"

**Supplementary Table 1. Characteristics of Age-group Classified Participants, Limited to Individuals without the *APOE*-ε2/ε4 Genotype.**

**A**

|  | SuperAgers | Middle-Aged  Controls | Old  Controls | Oldest-old  Controls | Middle-Aged  Cases | Old  Cases | Oldest-old  Cases |
| --- | --- | --- | --- | --- | --- | --- | --- |
| No. participants | 1393 | 1576 | 3128 | 1194 | 1091 | 3442 | 3166 |
| No. observations | 11389 | 6862 | 12004 | 6880 | 2673 | 10595 | 15032 |
| Visits, mean (SD) | 8 (5) | 4 (3) | 4 (3) | 6 (4) | 2 (2) | 3 (2) | 5 (4) |
| Follow-up time, mean (SD), y | 10 (6) | 6 (5) | 4 (4) | 8 (6) | 2 (2) | 2 (3) | 5 (6) |
| Baseline age, mean (SD), y | 77 (7) | 57 (5) | 70 (4) | 79 (7) | 59 (5) | 73 (4) | 83 (6) |
| Education, mean (SD), y | 16 (3) | 16 (2) | 16 (3) | 15 (3) | 15 (3) | 15 (3) | 15 (3) |
| Female Sex, No. (%) | 941 (68) | 1082 (69) | 1909 (61) | 651 (55) | 612 (56) | 1852 (54) | 1935 (61) |
| *APOE-*ε2 Frequency, No. (%) | 227 (16) | 170 (11) | 393 (13) | 160 (13) | 40 (4) | 98 (3) | 229 (7) |
| *APOE-*ε4 Frequency, No. (%) | 257 (18) | 601 (38) | 907 (29) | 251 (21) | 629 (58) | 2409 (70) | 1333 (42) |

**B**

|  | SuperAgers | Middle-Aged  Controls | Old  Controls | Oldest-old  Controls | Middle-Aged  Cases | Old  Cases | Oldest-old  Cases |
| --- | --- | --- | --- | --- | --- | --- | --- |
| No. participants | 200 | 276 | 720 | 140 | 82 | 392 | 425 |
| No. observations | 1729 | 942 | 2979 | 804 | 166 | 1225 | 2234 |
| Visits, mean (SD) | 9 (5) | 3 (3) | 4 (3) | 6 (4) | 2 (1) | 3 (3) | 5 (5) |
| Follow-up time, mean (SD), y | 9 (6) | 4 (4) | 4 (4) | 6 (5) | 1 (1) | 3 (3) | 5 (6) |
| Baseline age, mean (SD), y | 77 (6) | 60 (4) | 70 (4) | 79 (6) | 60 (4) | 73 (4) | 82 (6) |
| Education, mean (SD), y | 16 (3) | 15 (3) | 15 (3) | 13 (3) | 14 (3) | 14 (4) | 13 (4) |
| Female Sex, No. (%) | 168 (84) | 206 (75) | 540 (75) | 112 (80) | 57 (70) | 270 (69) | 314 (74) |
| *APOE-*ε2 Frequency, No. (%) | 45 (22) | 48 (17) | 123 (17) | 22 (16) | 5 (6) | 15 (4) | 34 (8) |
| *APOE-*ε4 Frequency, No. (%) | 43 (22) | 105 (38) | 229 (32) | 34 (24) | 62 (76) | 286 (73) | 197 (46) |

(A) Non-Hispanic White; (B) Non-Hispanic Black

**Supplementary Table 2. Comparisons of *APOE-*ε2 and *APOE*-ε4 Allele Frequencies among SuperAgers, Cases, and Controls, Limited to Individuals without the *APOE*-ε2/ε4 Genotype.**

**A**

|  | *APOE-*ε2 | | *APOE-*ε4 | |
| --- | --- | --- | --- | --- |
|  | OR (CI) | *P_FDR_* | OR (CI) | *P_FDR_* |
| SuperAgers vs. Middle-Aged Controls | 1.61 (1.30, 2.00) | *<0.001* | 0.37 (0.31, 0.43) | *<0.001* |
| SuperAgers vs. Old Controls | 1.37 (1.15, 1.64) | *0.001* | 0.55 (0.47, 0.65) | *<0.001* |
| SuperAgers vs. Oldest-Old Controls | 1.31 (1.04, 1.64) | *0.026* | 0.80 (0.66, 0.98) | *0.040* |
| SuperAgers vs. Middle-Aged Cases | 5.24 (3.68, 7.46) | *<0.001* | 0.17 (0.14, 0.20) | *<0.001* |
| SuperAgers vs. Old Cases | 7.24 (5.60, 9.37) | *<0.001* | 0.09 (0.08, 0.11) | *<0.001* |
| SuperAgers vs. Oldest-Old Cases | 2.62 (2.13, 3.21) | *<0.001* | 0.32 (0.27, 0.37) | *<0.001* |

**B**

|  | *APOE-*ε2 | | *APOE-*ε4 | |
| --- | --- | --- | --- | --- |
|  | OR (CI) | *P_FDR_* | OR (CI) | *P_FDR_* |
| SuperAgers vs. Middle-Aged Controls | 1.36 (0.86, 2.15) | 0.229 | 0.46 (0.30, 0.70) | *0.001* |
| SuperAgers vs. Old Controls | 1.44 (0.97, 2.11) | 0.093 | 0.60 (0.42, 0.88) | *0.016* |
| SuperAgers vs. Oldest-Old Controls | 1.46 (0.80, 2.66) | 0.251 | 1.03 (0.59, 1.78) | 0.947 |
| SuperAgers vs. Middle-Aged Cases | 4.36 (1.64, 11.61) | *0.007* | 0.10 (0.05, 0.18) | *<0.001* |
| SuperAgers vs. Old Cases | 8.13 (4.17, 15.84) | *<0.001* | 0.10 (0.07, 0.16) | *<0.001* |
| SuperAgers vs. Oldest-Old Cases | 3.38 (2.03, 5.62) | *<0.0001* | 0.34 (0.23, 0.51) | *<0.001* |

(A) Non-Hispanic White; (B) Non-Hispanic Black. Abbreviations: OR, Odds Ratio; CI, Confidence Interval (95%); *P_FDR_*, FDR-corrected P-value.

**Supplementary Table 3. Comparisons of *APOE-*ε2 and *APOE*-ε4 Allele Frequencies among SuperAgers, Cases, and Controls, with *APOE* genotyping method as an additional covariate.**

**A. Non-Hispanic White**

|  | *APOE-*ε2 | | *APOE-*ε4 | |
| --- | --- | --- | --- | --- |
|  | OR (CI) | *P_FDR_* | OR (CI) | *P_FDR_* |
| SuperAgers vs. Middle-Aged Controls | 1.39 (1.13, 1.71) | *0.002* | 0.38 (0.32, 0.45) | *<0.001* |
| SuperAgers vs. Old Controls | 1.24 (1.05, 1.47) | *0.018* | 0.56 (0.48, 0.65) | *<0.001* |
| SuperAgers vs. Oldest-Old Controls | 1.28 (1.03, 1.59) | *0.036* | 0.81 (0.66, 0.98) | *0.040* |
| SuperAgers vs. Middle-Aged Cases | 4.59 (3.31, 6.36) | *<0.001* | 0.18 (0.15, 0.22) | *<0.001* |
| SuperAgers vs. Old Cases | 4.04 (3.26, 5.00) | *<0.001* | 0.09 (0.08, 0.11) | *<0.001* |
| SuperAgers vs. Oldest-Old Cases | 2.03 (1.68, 2.45) | *<0.001* | 0.31 (0.27, 0.37) | *<0.001* |

**B. Non-Hispanic Black**

|  | *APOE-*ε2 | | *APOE-*ε4 | |
| --- | --- | --- | --- | --- |
|  | OR (CI) | *P_FDR_* | OR (CI) | *P_FDR_* |
| SuperAgers vs. Middle-Aged Controls | 1.16 (0.76, 1.78) | 0.549 | 0.49 (0.33, 0.73) | *0.001* |
| SuperAgers vs. Old Controls | 1.41 (0.99, 2.02) | 0.083 | 0.67 (0.47, 0.95) | *0.041* |
| SuperAgers vs. Oldest-Old Controls | 1.63 (0.93, 2.86) | 0.118 | 1.18 (0.70, 1.99) | 0.571 |
| SuperAgers vs. Middle-Aged Cases | 3.58 (1.58, 8.15) | *0.005* | 0.12 (0.07, 0.23) | *<0.001* |
| SuperAgers vs. Old Cases | 4.61 (2.77, 7.68) | *<0.001* | 0.13 (0.09, 0.19) | *<0.001* |
| SuperAgers vs. Oldest-Old Cases | 2.73 (1.76, 4.25) | *<0.001* | 0.39 (0.27, 0.57) | *<0.001* |

Abbreviations: OR, Odds Ratio; CI, Confidence Interval (95%); *P_FDR_*, FDR-corrected P-value.

NOTE: Counts for this comparison are unchanged from the main tables.

**Supplementary Table 4. Characteristics of Age-group Classified NHW Participants, Random Down-Sampling.**

|  | SuperAgers | Middle-Aged  Controls | Old  Controls | Oldest-old  Controls | Middle-Aged  Cases | Old  Cases | Oldest-old  Cases |
| --- | --- | --- | --- | --- | --- | --- | --- |
| No. participants | 211 | 289 | 750 | 145 | 84 | 413 | 444 |
| No. observations | 1805 | 1342 | 3189 | 726 | 222 | 1417 | 2033 |
| Visits, mean (SD) | 9 (6) | 5 (4) | 4 (3) | 5 (4) | 3 (2) | 3 (3) | 5 (5) |
| Follow-up time, mean (SD), y | 9 (6) | 6 (5) | 4 (4) | 5 (5) | 2 (2) | 3 (3) | 4 (5) |
| Baseline age, mean (SD), y | 78 (6) | 58 (5) | 70 (4) | 82 (6) | 59 (4) | 73 (4) | 84 (6) |
| Education, mean (SD), y | 17 (3) | 16 (2) | 16 (3) | 15 (3) | 15 (3) | 15 (3) | 15 (3) |
| Female Sex, No. (%) | 177 (84) | 216 (75) | 564 (75) | 115 (79) | 58 (69) | 283 (69) | 331 (75) |
| *APOE-*ε2 Frequency, No. (%) | 32 (15) | 38 (13) | 106 (14) | 23 (16) | 3 (4) | 17 (4) | 38 (9) |
| *APOE-*ε4 Frequency, No. (%) | 49 (23) | 105 (36) | 217 (29) | 32 (22) | 51 (61) | 310 (75) | 185 (42) |

**Supplementary Table 5. Comparisons of *APOE-*ε2 and *APOE*-ε4 Allele Frequencies among NHW SuperAgers, Cases, and Controls, Random Down-Sampling.**

|  | *APOE-*ε2 | | *APOE-*ε4 | |
| --- | --- | --- | --- | --- |
|  | OR (CI) | *P_FDR_* | OR (CI) | *P_FDR_* |
| SuperAgers vs. Middle-Aged Controls | 1.20 (0.72, 2.00) | 0.546 | 0.51 (0.34, 0.76) | *0.002* |
| SuperAgers vs. Old Controls | 1.12 (0.72, 1.72) | 0.657 | 0.73 (0.51, 1.04) | 0.089 |
| SuperAgers vs. Oldest-Old Controls | 1.02 (0.56, 1.87) | 0.950 | 1.08 (0.63, 1.84) | 0.777 |
| SuperAgers vs. Middle-Aged Cases | 5.54 (1.6, 19.16) | *0.011* | 0.20 (0.11, 0.35) | *<0.001* |
| SuperAgers vs. Old Cases | 4.54 (2.37, 8.69) | *<0.001* | 0.09 (0.06, 0.13) | *<0.001* |
| SuperAgers vs. Oldest-Old Cases | 2.08 (1.23, 3.52) | *0.011* | 0.41 (0.27, 0.60) | *<0.001* |

Abbreviations: OR, Odds Ratio; CI, Confidence Interval (95%); *P_FDR_*, FDR-corrected P-value.
